# Genetic Modifiers of Prenatal Brain Injury After Zika Virus Infection: A Scoping Review

**DOI:** 10.1101/2025.01.02.25319896

**Authors:** Fernanda J P Marques, Janet Ruan, Rozel B. Razal, Marcio Leyser, Youssef A. Kousa

## Abstract

**Introduction:** The severity of virally induced prenatal brain injury, even among dizygotic twins, varies according to individual and maternal risk and protective factors, including genomics.

**Objective:** This scoping review aims to analyze data on genetic susceptibility to neurological outcomes in children exposed in utero to Zika virus.

**Methods:** We followed JBI methodology for this scoping review. A search in PubMed, Scopus, CINAHL, Web of Science, Academic Search Ultimate, Agricola, Health Source: Nursing/Academic Edition, and Psychology and Behavioral Sciences Collection was conducted. Three reviewers independently screened studies using the Rayyan platform. Studies on gene mutations impacting brain injury after Zika virus infection were included.

**Results:** Thirteen articles identifying candidate genes related to brain injury were reviewed. Twenty-three genes were implicated in modulating susceptibility to prenatal brain injury, including six maternal and 17 infant genes.

**Conclusion:** Maternal and fetal genetic factors likely contribute susceptibility to virally induced prenatal brain injury. Analyzing polygenic risk could aid in future screening programs to identify individuals at risk. This information may eventually be integrated into clinical data, helping healthcare providers, families, and patients understand how to personalize care for better outcomes.

**Impact:**

- This paper evaluates available evidence about the relationship between genetic susceptibility and neurological consequences of Zika virus exposure during pregnancy.
- After performing a scoping review, we identified 13 articles describing candidate genes that potentially contribute to the development of virally induced brain injury after prenatal Zika infection. Of the genes identified, six were associated with maternal risks, while 17 were linked to the fetus.
- Maternal and prenatal genetic factors could increase the risk of virally induced prenatal brain injury.
- Future research should investigate factors that can modify disease pathogenesis toward the goal of reducing the global impact of brain injury.

## Introduction

In November 2015, the Brazilian Ministry of Health declared a state of public health emergency due to the rising number of Zika virus (ZIKV) cases in the country.^1^ Throughout 2015 and 2016, many pregnant people who experienced prenatal ZIKV infections gave birth to children with microcephaly, brain malformations, hearing loss, and locomotor development disability^2^. Since then, studies have also shown that ZIKV can lead to an increased risk of in-utero fetal death^3^.

In addition to associated comorbidities, the occurrence of severe brain malformations raised several concerns regarding child development. A novel clinical condition emerged, congenital Zika syndrome (CZS), which is associated with neurodevelopmental disability and early demise^4^. Many critical questions regarding CZS and its consequences remain unanswered, which could inform important gaps in our understanding of the natural history of this disease during pregnancy^5^.

Despite the epidemiologic and clinical significance of this problem, no prenatal standards of care in treatment are currently available, nor can virally induced prenatal brain injury be targeted for prevention. In addition, it is not clear why some infants are affected, and others are not. Differences in brain injury, even among dizygotic twins, suggest that there are genetic factors influencing viral infection and susceptibility to brain injury. Existing literature shows that specific genetic pathways can play a role in the development of brain injury from ZIKV^6^. This, along with the continued prevalence of the virus, warrants exploration of the virus’ corresponding neurobiology and genetic susceptibility as a critical step toward protecting the developing brain.

A growing body of evidence suggests that host genes can contribute to virally induced brain injury after ZIKV infection, but no comprehensive review has been conducted to compile this literature to date. A preliminary search of PubMed, MEDLINE, and JBI Evidence Synthesis was conducted and no current or ongoing systemic or scoping reviews on the topic were identified. A review of this topic is necessary to determine the range of information concerning genetic susceptibility for the development of virally induced brain injury and possible gaps in the literature for future studies. Such work can even one day inform the design of neuroprotective strategies.

The purpose of this scoping review is to identify candidate genes that are related to brain injury after prenatal ZIKV infection. Along with epidemiological, clinical, neuroimaging, and psychosocial risks, such information can be considered toward improving care for affected children.

## Methods

This scoping review considered studies related to possible candidate genes related to ZKV induced prenatal brain injury. Inclusion criteria were: experimental study designs that are randomized controlled trials, non-randomized controlled trials, before and after studies, and interrupted time-series studies. In addition, analytical observational studies, including prospective and retrospective cohort, case-control, and analytical cross-sectional studies were also considered for inclusion. We also included descriptive observational study designs, such as case series, individual case reports, and descriptive cross-sectional studies. Exclusion criteria were: gray literature, review and duplicated studies, and articles exclusively focused on host dependency or resistance factors associated with ZIKV infection but not related to ZIKV-related prenatal brain injury.

### Search Strategy

The search strategy aimed to locate both published and unpublished studies. The authors consulted with a public health librarian to design and refine the search and in developing a comprehensive search strategy and database selection for searches. The databases searched include PubMed, Scopus, Web of Science, Academic Search Ultimate, Agricola, Health Source: Nursing/Academic Edition, Psychology and Behavioral Sciences Collection, and CINAHL. An initial limited search of the Academic Search Ultimate, Agricola, Health Source: Nursing/Academic Edition, Psychology and Behavioral Sciences Collection was undertaken to identify articles on the topic also. Text in the titles and abstracts of relevant articles, and the index terms used to describe the articles, were used to develop a full search strategy.

The search strategy, including all identified keywords and index terms, was adapted for each included database and/or information source. The reference list of all included sources of evidence was screened for additional studies. Guidelines from the Preferred Reporting Items for Systematic Reviews and Meta-Analyses (PRISMA) and the Preferred Reporting Items for Systematic Review and Meta-Analysis Protocols (PRISMA-P) were applied to conduct the search (**Figure 1**). Studies published in any language were included. A third reviewer resolved any remaining disagreements or ambiguities.

**Figure. 1.**
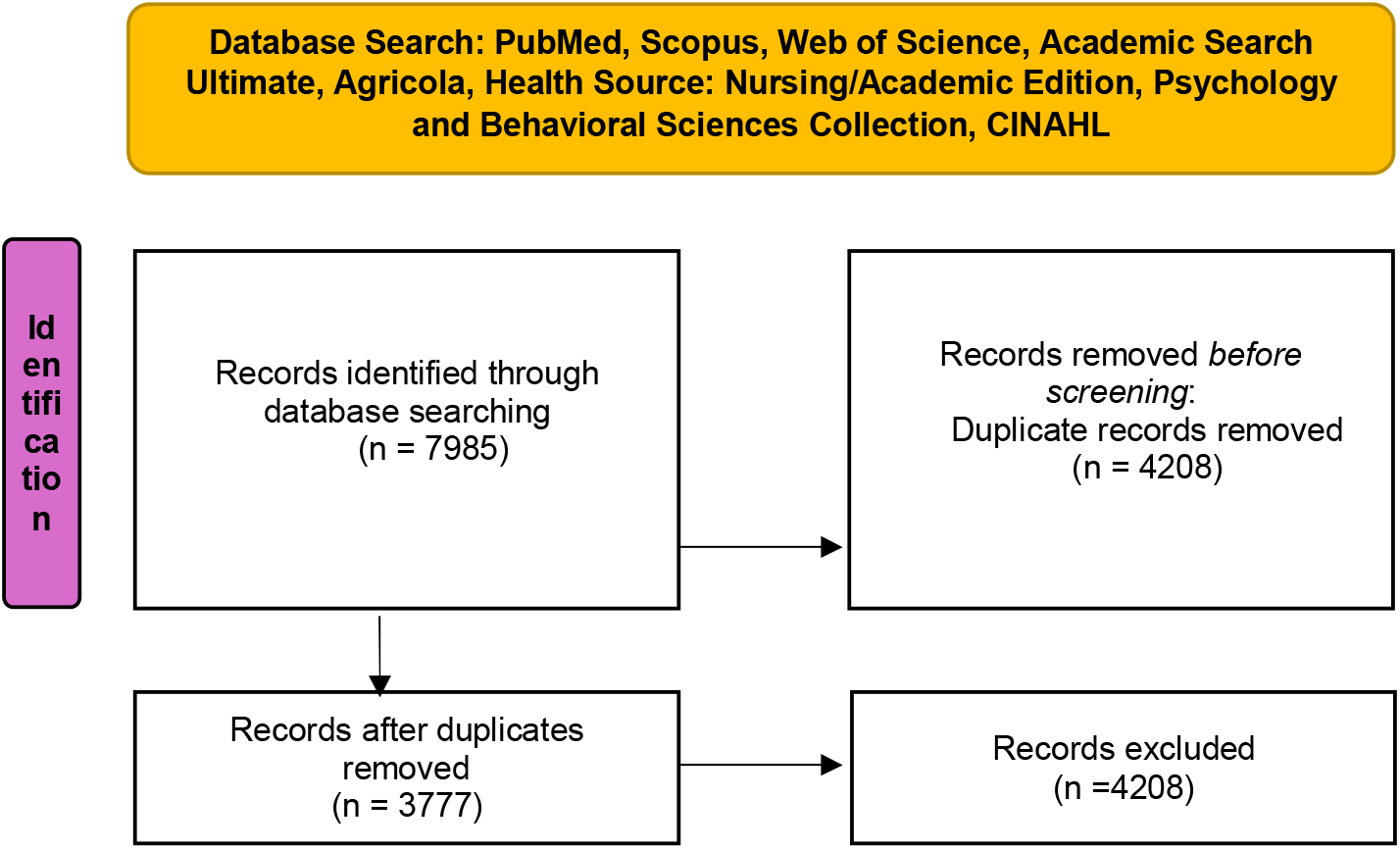
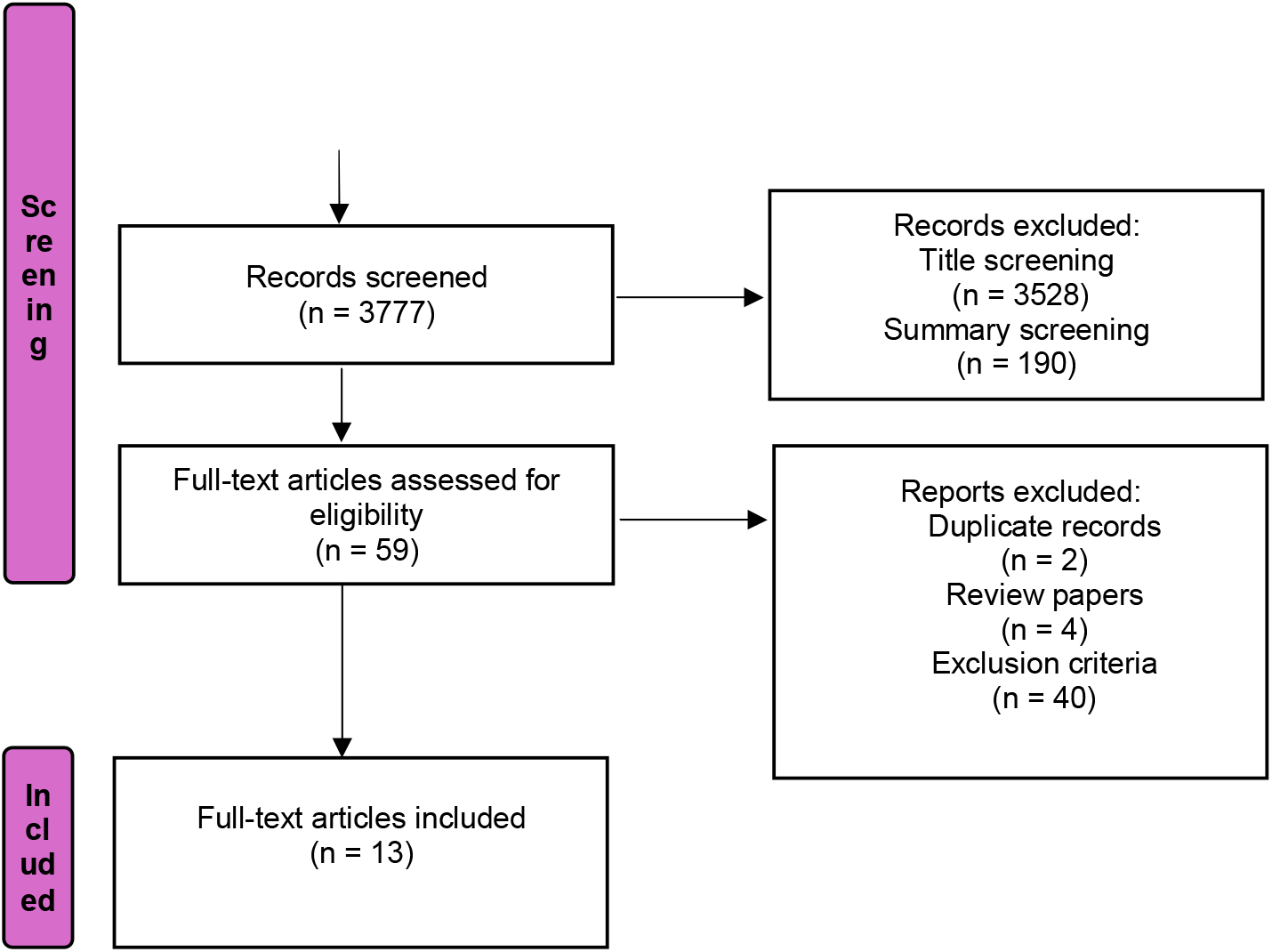
Scoping Review. PRISMA flow diagram. The PRISMA flow diagram for scoping review details the database searches, the number of abstracts screened, and the full texts retrieved.

The specifics included:

- <u>General Search Strategy:</u> Zika AND (“screen” OR “exome sequencing” OR “variant mutation” OR “disease susceptibility” OR “genetic susceptibility” OR “risk factors” OR “host factors” OR “host mutation” OR “microcephaly gene” OR “maternal genes” OR “infant genes” OR “susceptibility genes” OR mice* OR “human gene*” OR (Congenital AND syndrome AND (genetic*s* OR mutation)))
- <u>PubMed Search Strategy:</u> (“Zika Virus/genetics”[MeSH] OR “Zika Virus Infection/genetics*” [MeSH] OR “Antiviral Restriction Factors/genetics”[MeSH] OR “Zika Virus Infection/virology” [MeSH] OR Zika) AND (“screen” OR “exome sequencing” OR “Host-Derived Cellular Factors/genetics”[MeSH] OR “variant mutation” OR “disease susceptibility” OR “genetic susceptibility” OR “Polymorphism, Single Nucleotide/genetics”[MeSH] OR “risk factors” OR “host factors” OR “host mutation” OR “microcephaly gene” OR “maternal genes” OR “infant genes”?OR “susceptibility genes” OR mice* OR “human gene*” OR “Microcephaly/genetics” [MeSH] OR (Congenital AND syndrome AND (genetic*s* OR mutation)))

### Study/Source of Evidence Selection

Following the search, all identified citations were uploaded into the Rayyan platform for screening purposes. Following a pilot test, titles and abstracts were screened by three independent reviewers for assessment against the inclusion criteria for the review. Potentially relevant sources were marked in the Rayyan software. The full texts of the selected citations were assessed in detail against the inclusion criteria by three independent reviewers. The protocol of this review is published at the Open Science Framework (OSF) registries.

## Results

Corresponding to the inclusion criteria established for this scoping review, we found 13 articles (7-19) describing candidate genes that potentially contribute to virally induced brain injury after ZIKV infection. The PRISMA flowchart is detailed in **Figure 1**. The studies are summarized in **Table 1**.

**Table 1:** Full-text articles included in the scoping review.

| Author | Date | Genetic Factor | Summary |
| --- | --- | --- | --- |
| Amaral et al.<sup>7</sup> | 2020 | — | An RNA-seq analysis of hiPSC-derived trophoblasts from three pairs of dizygotic twins discordant for CZS was performed. However, potential viral attachment factors, including IFN genes and IFN receptor genes, were not differentially expressed. Global gene expression changes showed upregulation of *CDRT4* and *TMEM176A* and downregulation of *TGFB2-OT1* and *HIST2H4B* in the twin affected by CZS, but overall global gene expression levels were the same. |
| Borda et al.<sup>8</sup> | 2021 | *DISP3* rs2076469,<br>*IL12RB2* | This study included whole-exome sequencing and analysis of 40 unrelated infants exposed to prenatal ZIKV infection. In addition to the common variants in |
|  |  |  | *DISP3* and *IL12RB2* genes, the authors found 38 rare SNPs in genes related to neurological development and immunity. |
| Caires-Junior et al.<sup>9</sup> | 2018 | *DDIT4L*, *FOXG1*, *LHX2* | RNA-Seq of neural progenitor cells from pairs of twins discordant for CZS was performed. The genes *DDIT4L*, *FOXG1*, and *LHX2* showed the most significant difference, with lower expression levels among the affected twins. |
| Candelo et al.<sup>10</sup> | 2021 | *CDK5RAP2* | Genetic testing of an infant with CZS identified a mutation in *CDK5RAP2*. Prior research showed that diallelic variants of *CDK5RAP2* are linked with microcephaly. |
| Gomes et al.<sup>11, 12, 13</sup> | 2021, 2023 | *miR-124-1* rs531564<br>CG, TP53 rs1042522, *NOS2* rs2297518, *TNF* rs1799724 and haplotypes with higher TNF expression, *VEGFA* haplotypes | Among 88 infants exposed to ZIKV during pregnancy, genetic variants were evaluated in a series of candidate genes, including *VEGFA*, *PTGS2*, *NOS3*, *TNF*, *NOS2*, *TP53*, *MDM2*, *MIR605*, *LIF*, *miR-124* and *TFRC* genes. Individuals with CZS had a higher frequency of *NOS2* rs2297518 and the CG rs531564 polymorphism in *miR-124-1*. There were statistically significant differences in haplotype frequencies in *VEGFA*. The *TNF* rs1799724 allele was higher in children with CZS and severe microcephaly than in other children with CZS alone. There was also a difference among individuals with CZS and lissencephaly, who appeared to have a lower frequency of the G-allele in *TP53* rs1042522. |
| de O da Silva et al.<sup>14</sup> | 2021 | *MTOR* rs2295079 | An exome sequencing study of dizygotic twins with history of prenatal ZIKV infection included one infant with CZS. The team found 15 discordant alleles, and 11 could affect *MTOR signaling*. In particular, the rs2295079 variant was identified as likely causal. |
| Gomes et al.<sup>15</sup> | 2023 | — | A candidate gene approach was taken in evaluating 40 children exposed to ZIKV. The study did not identify a significant difference in AXL, TLR3, and STAT2 allele frequencies between children with and without CZS. |
| Rossi et al.<sup>16</sup> | 2019 | Maternal: rs11676272<br>*ADCY3*, *ADCY7* | A cohort of 52 women infected with ZIKV during pregnancy were evaluated for genetic risks in 205 candidate genes. The study identified 817 SNPs, possibly affecting the genes *ADCY3* and *ADCY7*. |
| Rossi et al.<sup>17</sup> | 2021 | Maternal: IL28B G allele<br>rs8099917 | In a follow-up study, the cohort of 52 women identified above were genotyped for seven Interferon-lambda non-coding single nucleotide polymorphisms (SNPs). A G-allele at rs8099917 showed an increased risk for the infant in having a diagnosis of CZS. |
| Santos et al.<sup>18</sup> | 2019 | Maternal: *TLR3*<br>rs3775291 Infant: *TNFA*<br>rs1799964 | Using a trio design (mother, father, and affected infant) along with unaffected participants, SNPs were evaluated among candidate genes. An association was found between a maternal SNP in *TLR3* (rs3775291) and the risk of the infant developing CZS. There was also an association between an infant having a SNP in *TNFA* (rs1799964) and the occurrence of severe microcephaly. |
| Santos et al.<sup>19</sup> | 2023 | Maternal: C allele<br>*TREM1* rs2234246, C<br>allele *IL4* rs224325.<br>Infant: *CXCL8* rs4073,<br>G allele *CXCL10*<br>rs4508917, *CXCL8*<br>rs4073, *TRL7* rs179008 | Using a case-control design, mother and infants affected or not affect by CZS were evaluated for SNPs in candidate genes. Genes evaluated included *TREM1*, *CXCL10*, *IL4*, *CXCL8*, *TLR3*, *TLR7*, *IFNR1*, *CXCR1*, *IL10*, *CCR2* and *CCR5*. SNPs in *TREM1* and *IL4* in the mother were associated with increased risk of CZS in infants. |

Caption: Thirteen articles describing candidate genes that potentially contribute to the development of CZS, affecting either the mother or the fetus.

Our scoping review has identified 23 genes that might have potential roles in modulating susceptibility to virally induced brain injury after ZIKV infection, including CZS. These included six maternal and 17 fetal/neonatal genes, that sometimes were associated with injury and at other times with protection.

Since CZS impacts the developing brain, it is reasonable that candidate gene approaches focused on genes and related pathways required for brain development. These included DISP3, miR-124-1, TP53, NOS2, VEGFA, MTOR, CDK5RAP2, FOXG1, and LHX2, which contribute to neurogenesis, neuronal differentiation, angiogenesis, microtubule nucleation, cell cycle regulation, and centriole attachment^8,9,10,11,12,13,14^. Other candidate genes approaches evaluated inflammation and immune response as the context for brain injury is a prenatal viral infections. Such genes included IL12RB2, CXCL8, CXCL10, among other interleukin-related genes^8,19^. Additional candidates evaluated included VEGFA, TNF, and NOS2, which are also associated with immune and inflammatory regulation during viral replication^11,12,13^. Related, IL12RB2 and TRL7 also play roles in innate and adaptive immunity^8,11,12,13,19^. In the cellular response, MTOR and DDIT4L are related to the endoplasmic reticulum, autophagy, and cellular stress^9,14^. The six identified maternal genes (TREM1, IL4, ADCY3, ADCY7, IL28B, TLR3) are also related to immune responses^16,17,18,19^. Though we encountered genes associated with ZIKV infection, the only genes included in this review are those that contribute specifically to the pathogenesis of virally induced brain injury^20,21,22^.

## Discussion

While there is data to support both maternal and fetal genetic modifiers in prenatal brain injury after ZIKV infection, there are several limitations that need to be addressed in future studies. For instance, as this is a rare disorder, cohorts in these studies tended to be smaller. To limit the impact of multiple testing in association studies, a candidate gene approach was taken. Future replication studies on these candidate genes or genome wide will require larger cohorts to account for population stratification.

Further, studies investigating pathogenic variants that modify susceptibility to ZIKV infection often compare infected individuals to healthy controls. In contrast, studies focusing on CZS can evaluate for specific risks within the infected group, comparing those affected with CZS to those affected without CZS. This narrows the available subjects for CZS studies, making it difficult to identify a genetic risk factor that is unique to virally induced brain injury. Therefore, future studies might consider pooling more samples or phenotypes for larger genomic association studies.

Although current research findings do not point to a monogenic model to explain CZS, it is important to conduct functional analyses on the candidate genes that were listed ^9^. Gene overexpression or gene knockout experiments using cellular or animal models would validate the role of these genes in the pathogenesis of CZS and could identify cellular pathways in developing therapies in the future.

This review focused on severe brain injury and CZS, which is characterized by microcephaly and other brain anomalies in infants prenatally exposed to ZIKV infection^3^. Other pathogens, such as Cytomegalovirus (CMV) and herpes simplex virus (HSV), can also cause virally induced prenatal brain injury. These viruses have been categorized as “teratogens” for the severe impact on the fetus that results far too often. Outcomes after such severe brain injury includes neurodevelopmental disorders such as cerebral palsy, autism spectrum disorder, and cognitive impairment^23^. Of interest, recent findings have also shown that prenatal exposure to SARS-CoV-2 can negatively affect brain development^24^. Since many of the candidate genes for CZS are involved in neurodevelopment, it would be interesting to investigate if these genes also contribute to the pathogenesis of other viral infections in future studies.

Many questions about ZIKV and its consequences remain unanswered, particularly why some infants exposed to ZIKV during pregnancy develop severe brain injury while others do not. Currently described markers for developmental disorders associated with ZIKV exposure (including microcephaly and calcifications) may appear late or may not be present at all, leading to a delay in surveillance and diagnosis^5^. However, the acute clinical, neurological, and pathophysiological features of severe brain injury frequently lead to life-long disabilities that affect not only impact the patients but also their families and communities. Therefore, it is essential to investigate factors specific to CZS early in its disease pathogenesis to reduce its overall impact on health, social structures, and economics.

This includes identifying host genetic risk or protective factors that can explain why there are difference in outcome after prenatal exposure to ZIKV and other viral infections. Such work may even one day inform our understanding of the connection between viral infection and brain injury. The design of a pipeline based on multilevel research might result in predictive models that would likely be beneficial for early identification of newborns and infants at risk for severe brain injury or CZS^25^. The approach might be multifaceted, designed to include epidemiological, clinical, neuroimaging, laboratory, genetics, neurophysiology, developmental, and psychosocial factors that, together, form the still-enigmatic chronic stages of the CZS’s biopsychosocial complex.

## Data Availability

All data produced in the present work are contained in the manuscript

## Author Contributions

FJPM conceptualized and designed the study, coordinated and supervised data collection, and drafted, reviewed, and revised the manuscript. JR contributed to study design, data collection instruments, provided review and analyses of data, and reviewed and revised the manuscript. RR contributed to data collection instruments, provided review and analyses of data, and reviewed and revised the manuscript. ML and YAK conceptualized and contributed to the design of the study and early drafts. ML and YAK critically reviewed the manuscript for important intellectual content. YAK supervised the project. All authors approved the final manuscript as submitted and agree to be accountable for the work.

## Acknowledgements

We thank Ms. Nedelina Tchangalova, public health librarian, for refining the search strategy and database selection. We also thank Ms. Zoe Kang for her critical review and feedback on the manuscript. We are grateful to support from our institutions and networks, including the SARAH Network, Rio de Janeiro, Brazil, Children’s National Research Institute in Washington, D.C., University of Iowa, Iowa City, and the PING Consortium.

## Author Information

